# Early statin use is associated with improved survival and cardiovascular outcomes in patients with atrial fibrillation and recent ischaemic stroke

**DOI:** 10.1101/2024.05.21.24307724

**Authors:** Sylvia E. Choi, Tommaso Bucci, Jia-Yi Huang, Kai-Hang Yiu, Christopher T. W. Tsang, Kui Kai Lau, Andrew Hill, Greg Irving, Gregory Y. H. Lip, Azmil H. Abdul-Rahim

## Abstract

**Background:** Statins reduce recurrent stroke and cardiovascular events in patients with non-cardioembolic stroke. We aimed to investigate the benefits of statins in patients with atrial fibrillation (AF) and recent ischaemic stroke (IS), which remain unclear.

**Methods:** This retrospective, cohort study was conducted using deidentified electronic medical records within TriNetX platform. Patients with AF and recent IS, who received statins within 28 days of their index stroke were propensity score-matched with those who did not. Patients were followed-up for 2 years. Primary outcomes were the 2-year risk of recurrent IS, all-cause mortality and the composite outcome of all-cause mortality, recurrent IS, transient ischaemic attack (TIA), and acute myocardial infarction (MI). Secondary outcomes were the 2-year risk of TIA, intracranial haemorrhage (ICH), acute MI, and hospital readmission. Cox regression analyses were used to calculate hazard ratios (HRs) with 95% confidence intervals (95%CI).

**Results:** Of 20,902 patients with AF and recent IS, 7,500 (35.9%) received statins within 28 days of their stroke and 13,402 (64.1%) did not. 11,182 patients (mean age 73.7±11.5; 5,277 (47.2%) female) remained after propensity score matching. Patients who received early statins had significantly lower risk of recurrent IS (HR: 0.45, 95%CI: 0.41–0.48, p<0.001), mortality (HR: 0.75, 95%CI: 0.66-0.84, p<0.001), the composite outcome (HR: 0.48, 95%CI: 0.45-0.52, p<0.001), TIA (HR: 0.37, 95%CI: 0.30-0.44, p<0.001), ICH (HR: 0.59, 95%CI: 0.47-0.72, p<0.001), acute MI (HR: 0.35, 95%CI: 0.30-0.42, p<0.001) and hospital readmission (HR: 0.46, 95%CI: 0.42-0.50, <0.001). Beneficial effects of early statins were evident in the elderly, different ethnicities, statin dose intensity, and AF subtypes, large vessel occlusion and embolic strokes and within the context of statin lipophilicity and optimal LDL-C levels.

**Conclusions:** Patients with AF and recent IS, who received early statins, had a lower risk of recurrent stroke, death, and other cardiovascular outcomes including ICH, compared to those who did not.

## Introduction

After ischaemic heart disease, stroke is the world’s second leading cause of death and third leading cause of disability^1^. Atrial fibrillation (AF) is a major cause of cardioembolic stroke present in up to 25% of all IS, which are often more severe or fatal^2,3^. The recurrence of IS is high, up to 35% at 5 years and up to 50% at 10 years^4^. The risk of recurrent stroke in non-anticoagulated patients with AF and history of previous stroke is 2.5 fold higher, compared to anticoagulated patients, with an average rate of 10% yearly^5,6^. This is particularly important given the multimorbidity burden associated with such patients^7,8^, necessitating a holistic or integrated care approach to post-stroke management to mitigate clinical cardiovascular outcomes^9,10^.

Evidence for the beneficial effects of statin therapy in patients with a history of coronary heart disease (CHD) or at high risk of cardiovascular events is well documented^11,12^. Moreover, the use of statins for prevention of recurrent strokes in non-AF related and non-cardioembolic IS patients without a history of CHD is well established^13–16^. However, many major secondary prevention trials excluded patients with AF or patients who had cardioembolic strokes. In addition, whether the perceived higher risks of haemorrhagic strokes associated with statin use observed in some trials would extend to such patients, or would be influenced by stroke sub-type, is unknown. Thus, there remains a gap in evidence to support the role of early statin use in AF-related stroke. No specific recommendations on the use of statins for secondary prevention in cardioembolic stroke exists in clinical practice guidelines.

On the basis of current clinical evidence, many international stroke guidelines on the secondary prevention of stroke recommend intensive statin therapy for stroke patients with non-cardioembolic strokes or history of atherosclerotic disease, or those at very high-risk^17,18^. The European Society of Cardiology (ESC) guidelines on management of dyslipidaemia and the European Stroke Organisation (ESO) guidelines on long-term secondary prevention after IS^19,20^, go even further. As patients with a history of IS are considered at very high risk of atherosclerotic cardiovascular disease, particularly recurrent IS, such patients should receive intensive lipid lowering therapy^19,20^.

Nevertheless, knowing whether or not to initiate statins in patients with AF and recent IS would help to clarify a familiar management conundrum. In this study, we aimed to investigate the effect of early statin therapy on clinical outcomes in patients with AF who have recently experienced an IS using a global federated research network.

## Methods

A request for access to TriNetX data can be made (https://live.trinetx.com). A data sharing agreement would be required, costs may be involved, and only de-identified information would be available.

### Data Source

The study used TriNetX, a global federated health research network containing electronic medical records (EMR) data provided by participating healthcare organisations (HCOs). HCOs are typically academic health centre-based healthcare systems or non-academic research-focused HCOs, and include hospitals, specialist physician practices, and primary care providers. Although HCOs are mainly within the United States, the network extends to 30 countries in North and South America, Europe, the Middle East, Africa, and the Asia-Pacific regions^21^. Data including demographics, encounters, diagnoses (using International Classification of Diseases, Tenth Revision, Clinical Modification [ICD-10-CM] codes), procedures, medications, laboratory results, and vital signs are accessible from 130 HCOs covering 170 million patients^21^.

TriNetX is compliant with applicable regional data protection, security and privacy laws and regulations, such as the Health Insurance Portability and Accountability Act (HIPAA) in the United States, and the General Data Protection Regulation (GDPR) in Europe^21^. Participating HCOs in the TriNetX network provide EMR data in de-identified, pseudo-anonymised, or limited form. The identity of each HCO remains anonymous and the data they share through the TriNetX Platform are attenuated to ensure that they do not include sufficient information to facilitate the determination of which HCO contributed specific information about a patient. To safeguard against the risk of patient reidentification, the TriNetX platform only uses aggregated counts and statistical summaries of de-identified information. As no patient identifiable information is received, studies using the TriNetX network do not require ethical approval or informed consent. Data is de-identified according to the de-identification standard defined in Section §164.514(a) of the HIPAA Privacy Rule. In the United States, the process by which data is de-identified has been attested to through a formal determination by a qualified expert as defined in Section §164.514(b)(1) of the HIPAA Privacy Rule. This formal determination by a qualified expert refreshed on December 2020.

### Study design

This was a retrospective observational study using complete case, de-identified data from the TriNetX platform. The data used in this study was collected on 11^th^ December, 2023 from the TriNetX Global Collaborative Network, which provided access to EMR from approximately 106 million patients aged 18 or over from over 110 healthcare organizations in up to 16 countries. This study followed the Strengthening the Reporting of Observational Studies in Epidemiology (STROBE) reporting guideline for observational studies.

### Cohort

Patients aged 18 or over with AF or flutter (ICD-10-CM code I48) and recent IS (ICD-10-CM code I63 (Cerebral infarction)) recorded in EMRs between 4^th^ May 2008 and 3^rd^ May 2018 were identified (**Table S1**). At the time of the search, a total of 106 participating HCOs responded including HCOs returning data on the platform for patients who met the study inclusion criteria. Patients were excluded if they had received statin treatment within the year leading up to the date of the index stroke. The baseline index event date was the date that a patient was diagnosed with IS for the first time.

The cohort was divided into two groups using electronic health records: (i) individuals who received treatment with statins within 28 days of their index stroke; and (ii) individuals who did not receive statins within 28 days of their index stroke.

To mitigate the risk of time-related biases, the follow-up period started 28 days after the index stroke only in those who were alive at that date, and individuals were included in the study if they received either angiotensin converting enzyme inhibitors or angiotensin II inhibitors within 28 days of their index stroke.

### Outcomes

Patients who received treatment with statins within 28 days of their index stroke were propensity score matched with patients who did not, before comparing outcomes between the two groups. All patients were followed-up for up to 2 years. Primary outcomes were the 2-year risk of recurrent IS (ICD-10-CM code I63), all-cause mortality and the composite outcome of recurrent IS, transient ischaemic attack (TIA) (ICD-10-CM code G45), acute myocardial infarction (MI) (ICD-10-CM code I21) and all-cause mortality (**Table S1**).

Secondary outcomes were the 2-year risk of TIA, intracranial haemorrhage (ICH) (ICD-10-CM code I61-I62), acute MI, and hospital readmission (**Table S1**).

### Statistical analysis

All statistical analyses were performed on the TriNetX online research platform. Baseline characteristics were compared using chi-squared tests for categorical variables and independent-sample t-tests for continuous variables. Propensity score matching was used to control the differences in the comparison cohorts. Cohort matching was performed for age at index stroke, gender, ethnicity, baseline comorbidities (hypertension, ischaemic heart diseases, peripheral vascular diseases, heart failure, lipid disorders, type 2 diabetes mellitus, chronic kidney disease, neoplasms, overweight and obesity, pulmonary heart disease, and obstructive sleep apnoea), medications (β-blockers, diuretics, antiarrhythmics, calcium channel blockers, angiotensin converting enzyme inhibitors, angiotensin II inhibitors, prior use of antiplatelet agents or anticoagulants, antianginal medications, insulin and oral hypoglycaemic agents) and cardiac procedures (cardiography, echocardiography, cardiac catheterisation, elective DC cardioversion, pacemaker and implanter defibrillator procedures, and intracardiac electrophysiological procedures) (**Table S1**). Those variables were selected because they may influence the clinical outcomes.

TriNetX performs a 1:1 greedy nearest neighbour matching model with a caliper of 0.1 pooled standard deviations using logistic regression. Any baseline characteristic with a standardised mean difference between cohorts lower than 0.1 is considered well matched. After propensity score matching, Cox-regression proportional hazard models were used to calculate hazard ratios with 95% confidence intervals to assess the association between treatment with statins within 28 days of an index stroke and the 2-year incidence of primary and secondary outcomes. The proportional hazards assumption was assessed using a scaled Schoenfeld residual-based approach. Differences in Kaplan-Meier curves between the group that received early statins and the group that did not were evaluated with the log-rank test. Patients were censored from the Kaplan-Meier analysis at the end of the follow-up period or on the day after the last entry in their record (if earlier). A patient having an outcome after their index stroke and on or before the date falling 28 days after their index stroke, was censored at the end of the 28-day period. To test the generalisability of our hypothesis, we conducted two sensitivity analyses to investigate the risk of the primary outcomes: (i) in older patients aged ≥75 years; and (ii) without propensity score matching. We conducted subgroup analyses examining the effects of early statins across different ethnicities, statin dose intensities, LDL-C levels, and within the context of statin lipophilicity, large vessel occlusion and embolic strokes, and different AF subtypes. All analyses were performed in the TriNetX platform which uses R’s survival package v3.3. The level of statistical significance was set at p<0.05.

## Results

### Cohort Characteristics

We identified 20,902 individuals who met the study inclusion criteria. Of these, 7,500 (35.9%) received statins within 28 days of their index stroke, and the remaining 13,402 (64.1%) did not receive statins within 28 days of their stroke. **Table S2** summarised the baseline characteristics of the two cohorts before and after propensity score matching, respectively.

Compared to patients who did not receive early statins, the group who received early statins had a statistically higher prevalence of white patients (69.8 vs 64.7%, p<0.001), and a lower prevalence of Black or African American patients (10.7 vs 11.9%, p=0.010) and female patients (46.6 vs 48.3%, p=0.019). Statin users were less likely to suffer from major comorbidities, and were more likely to receive cardiovascular medications (such as betablockers, angiotensin-converting enzyme inhibitors, calcium channel inhibitors and diuretics), antiplatelet agents, anticoagulants, insulin and oral medications for diabetes.

After propensity score matching, a total of 11,182 well-matched patients remained (mean age 73.7±11.5; 5,277 (47.2%) female). Each group (n=5,591) was followed up for two years for the primary and secondary outcomes.

### Primary outcomes: recurrent IS, all–cause mortality, and the composite outcome

After propensity score matching, compared with patients who did not receive early statins following their index stroke, those that did receive early statins had a significantly lower risk of all primary outcomes. Early statin use was associated with a significantly lower risk of recurrent IS within two years (872 (15.6%) vs. 2,130 (38.1%); Hazard Ratio [HR]: 0.45, 95% Confidence Interval [CI]: 0.41–0.48, p<0.001). The risk of all-cause mortality within two years was reduced (444 (8.0%) vs. 793 (14.2%); HR: 0.75, 95%CI: 0.66-0.84, p<0.001), as was the risk of the composite outcome (1,375 (24.6%) vs. 2,945 (52.7%); HR: 0.48, 95%CI: 0.45-0.52, p<0.001) in patients who received early statin therapy, compared with those who did not (**Figure 1**).

**Figure 1:**
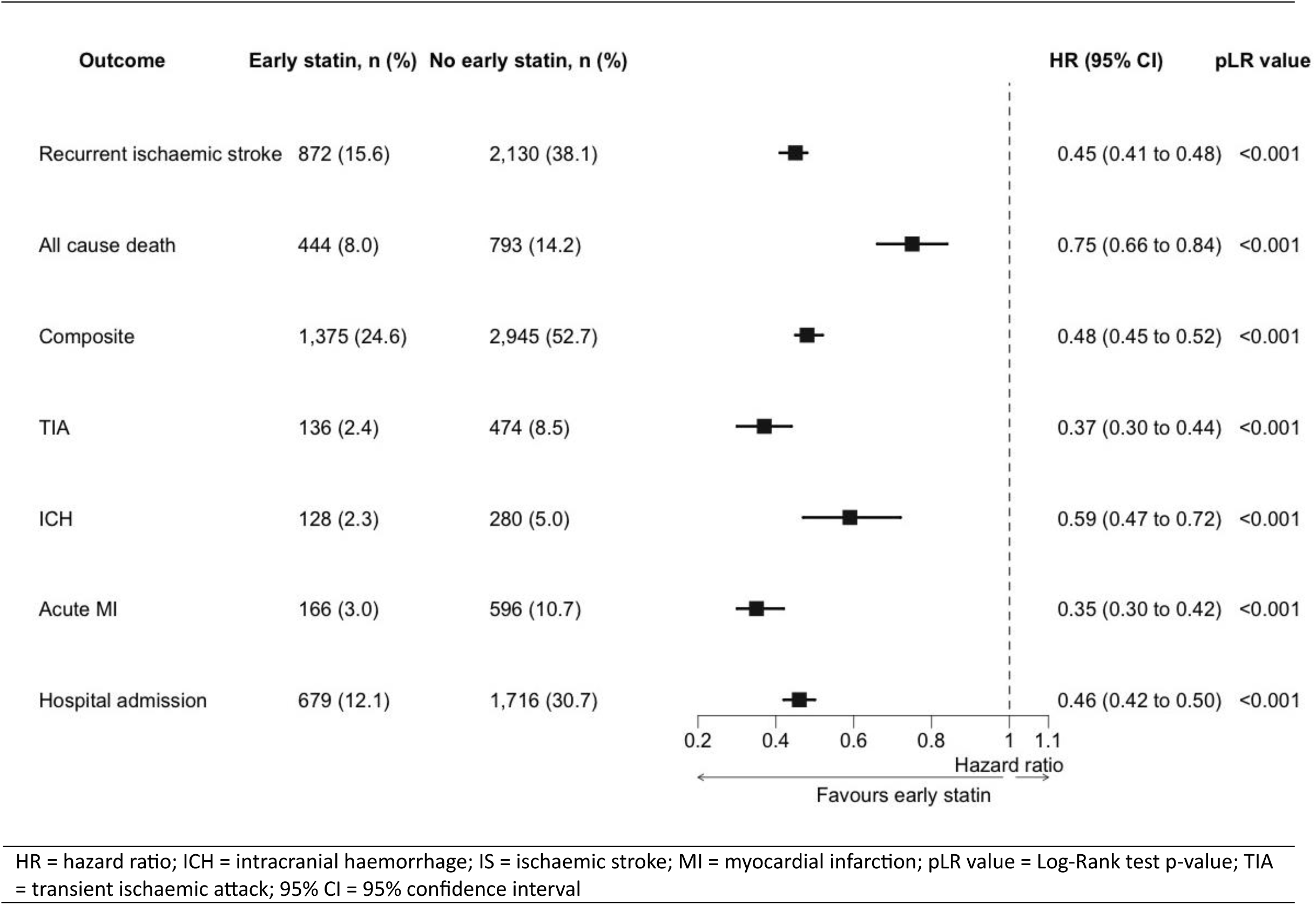
Cox regression survival analysis of primary and secondary outcomes

### Secondary Outcomes

Patients who received early statins within 28 days of their index stroke had a significantly lower risk of the secondary outcomes, compared to patients who did not receive early statins. Early statin use was associated with a lower risk of TIA (136 (2.4%) vs. 474 (8.5%); HR: 0.37, 95%CI: 0.30-0.44, p<0.001), ICH (128 (2.3%) vs. 280 (5.0%); HR: 0.59, 95%CI: 0.47-0.72, p<0.001), acute MI (166 (3.0%) vs. 596 (10.7%); HR: 0.35, 95%CI: 0.30-0.42, p<0.001) and hospital readmission (679 (12.1%) vs. 1,716 (30.7%); HR: 0.46, 95%CI: 0.42-0.50, p<0.001) (**Figure 1**).

### Sensitivity analyses and sub-group analyses

We conducted sensitivity analyses without adjusting for the effects of propensity score matching and found that results were similar for the primary outcomes. Before propensity score matching, early statin use was associated with a lower risk of recurrent IS (1,144 (15.2%) vs. 5,098 (38.0%); HR: 0.44, 95%CI: 0.41-0.47, p<0.001), all-cause mortality (593 (7.9%) vs. 2,002 (14.9%); HR: 0.70, 95%CI: 0.64-0.77, p<0.001), and the composite outcome (1,819 (24.2%) vs. 7,023 (52.4%); HR: 0.48, 95%CI: 0.46-0.51, p<0.001) after two years’ follow-up (**Figure 2**).

**Figure 2:**
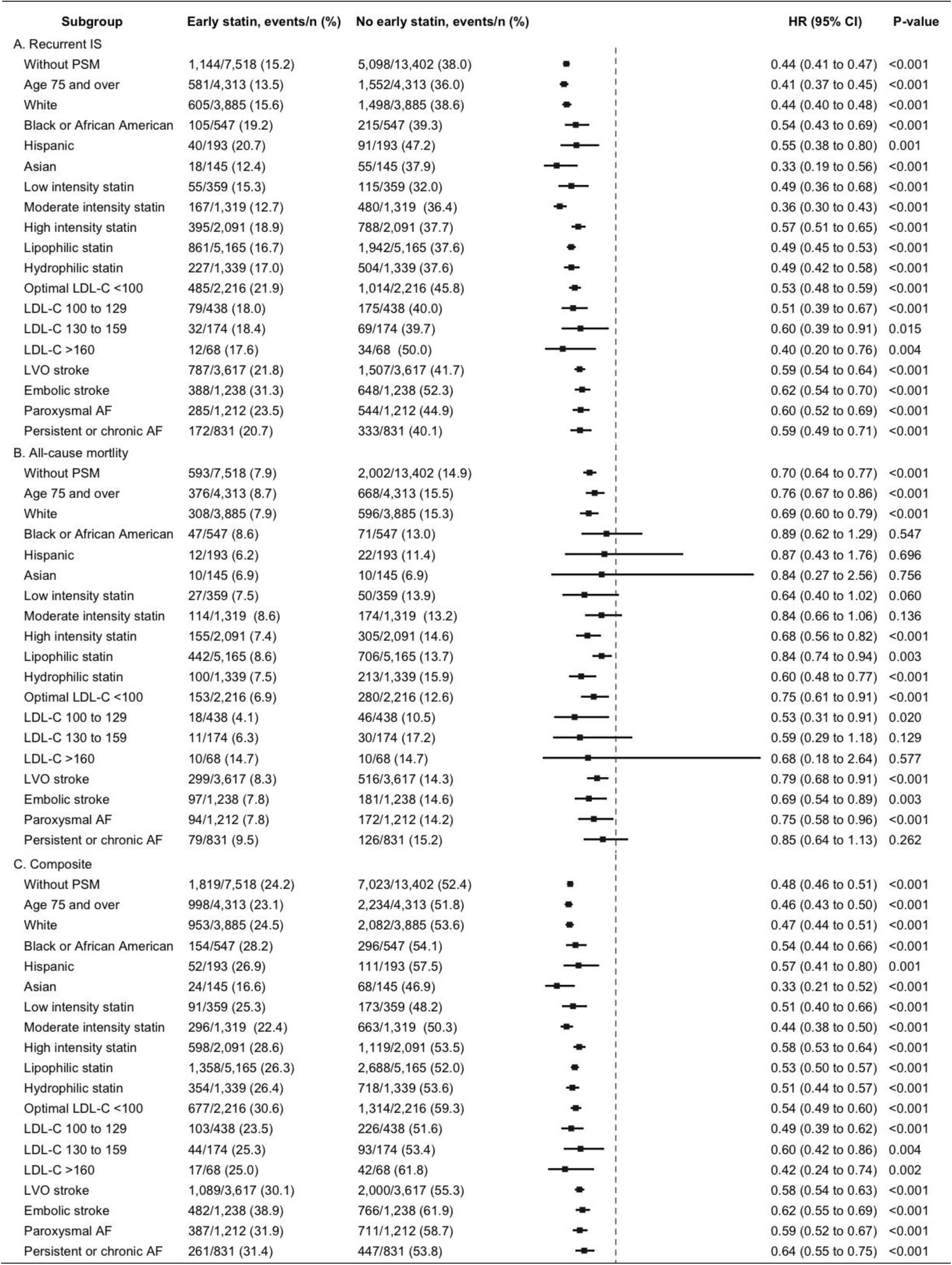
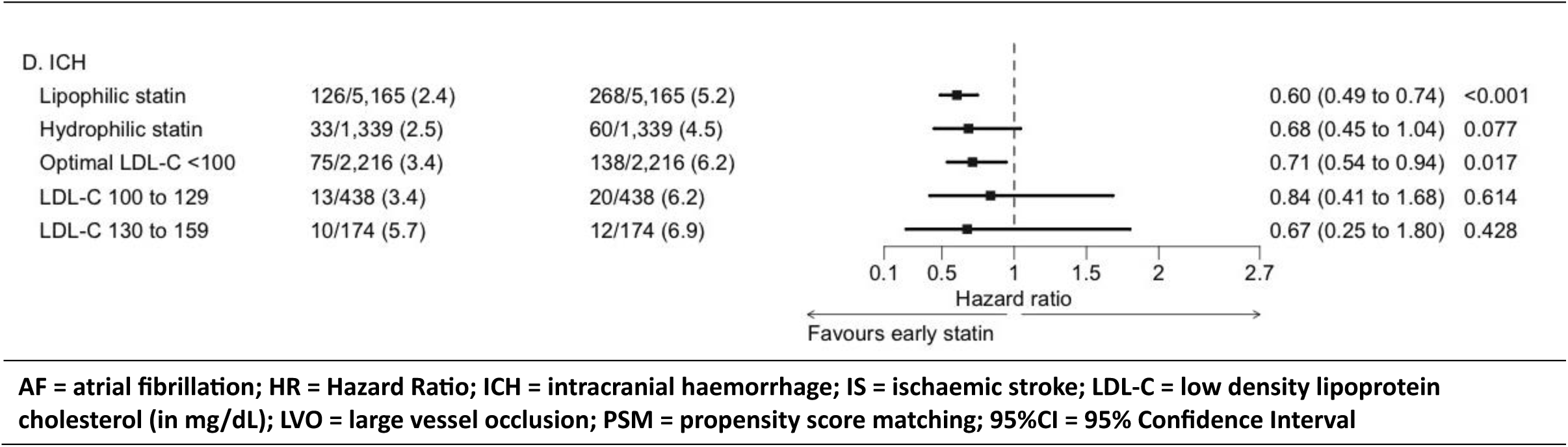
Sensitivity and subgroup analyses using Cox regression

The incidence of both stroke and AF increases with age. To evaluate the benefit of statins for secondary prevention of AF-related stroke in older patients, we conducted a sensitivity analysis for patients aged 75 years and over. We identified 15,840 patients aged ≥75 years with AF and recent IS (mean age 78.8 ± 7.2, 52.1% female), 5762 (36.4%) of whom received early statins, and 10,078 (63.4%) of whom did not. After propensity score matching, there were 4,313 well-matched individuals in each group. **Table S3** showed the baseline characteristics of individuals aged ≥75 years before and after propensity score matching.

Among patients aged ≥75 years, early statin use was associated with a significantly reduced risk of all primary outcomes within two years. There was a lower risk of recurrent IS (581 (13.5%) vs. 1,552 (36.0%); HR: 0.41, 95%CI: 0.37-0.45, p<0.001), all-cause mortality (376 (8.7%) vs. 668 (15.5%); HR: 0.76, 95%CI: 0.67-0.86, p<0.001), and the composite outcome (998 (23.1%) vs. 2,234 (51.8%); HR: 0.46, 95%CI: 0.43-0.50, p<0.001) (**Figure 2**).

### Ethnicity

Patients falling within the White ethnic group who received early statins showed reductions in the risk of all primary outcomes, similar to the main analysis (**Figure 2**). Patients in the Black or African American ethnic group who received early statins were associated with a lower risk of recurrent stroke and of the composite outcome, compared to those who did not receive early statins. Due to the small size of that ethnic group, no statistical difference could be established in all-cause mortality between the early statin and no-early statin groups and the 95%CIs were wide (HR: 0.89, 95%CI: 0.62-1.29) (**Figure 2**). Findings similar to those reported in the Black or African American ethnic group were observed in the Hispanic and Asian ethnic groups (**Figure 2**).

### Dose intensity of statin

We compared three subgroups comprising patients receiving (1) low-intensity statin (2) moderate-intensity statin and (3) high-intensity statin (in each case, as defined by the American College of Cardiology/American Heart Association Guideline on the Management of Blood Cholesterol)^22^.

After propensity score matching, compared to patients who did not receive early statins, patients in the high-intensity statin sub-group showed a lower risk of all primary outcomes, with a reduction in the risk of recurrent stroke (395 (18.9%) vs. 788 (37.7%); HR: 0.57, 95%CI: 0.51-0.65, p<0.001), all-cause mortality (155 (7.4%) vs. 305 (14.6%); HR: 0.68, 95%CI: 0.56-0.82, p<0.001), and the composite outcome (598 (28.6%) vs. 1,119 (53.5%); HR: 0.58, 95%CI: 0.53-0.64, p<0.001). Patients in the low-intensity and moderate-intensity statins subgroups showed a reduction in the risk of recurrent stroke and the composite outcome; however, there was no difference in the risk of all-cause mortality between the no-early statin groups and the low-intensity statin or moderate intensity statin sub-groups (**Figure 2**).

### Lipophilicity of statins

We analysed two subgroups including (1) patients receiving early statins regarded as relatively lipophilic (atorvastatin, simvastatin, fluvastatin, lovastatin, pitavastatin, and cerivastatin), and (2) patients receiving early statins regarded as relatively hydrophilic (pravastatin and rosuvastatin).

After propensity score matching, compared to patients who did not receive early statins, patients in each subgroup receiving either lipophilic or hydrophilic statins, had significantly lower risk of all primary outcomes (**Figure 2**). There was a reduced risk of ICH in the lipophilic statin group when compared to the no-early statin group (HR 0.60, 95%CI: 0.49-0.74). This difference was not seen when comparing the hydrophilic statin group with the no-early statin group (HR 0.68, 95%CI: 0.45-1.04) (**Figure 2**).

### LDL-C Level

Compared to patients with optimal low-density lipoprotein cholesterol (LDL-C) who did not receive early statins, the subgroup of patients with optimal LDL-C who did receive early statins had a lower risk of all three primary outcomes and a reduced risk of ICH (**Figure 2**).

### Large vessel occlusion and embolic stroke

Two subgroup analyses of AF patients with recent large vessel occlusion (LVO) IS (ICD-10-CM code I63.5 (Cerebral infarction due to unspecified occlusion or stenosis of cerebral arteries)) and AF patients with recent embolic strokes (ICD-10-CM code I63.4 (Cerebral infarction due to embolism of unspecified cerebral artery)), demonstrated an association between early statin use and a lower risk of all primary outcomes after two-year follow-up (**Figure 2**).

### AF subtype

We conducted two subgroup analyses of (1) patients with persistent and chronic AF and recent IS (ICD-10-CM code 148.1 and 148.2) and (2) patients with paroxysmal AF (ICD-10-CM code 148.0) and recent IS. After propensity score matching, the subgroup of patients with paroxysmal AF demonstrated an association between early statin use and a lower risk of all primary outcomes (**Figure 2**).

A smaller subgroup of patients with persistent and chronic AF showed an association between early statin use and a reduced risk of recurrent IS and the composite outcome (HR 0.64, 95%CI: 0.55-0.75). There was no significant difference between the early statins group and the no-early statin group with regards to the endpoint of all-cause mortality.

## Discussion

In this retrospective, propensity-score matched analysis of 11,182 individuals with AF and recent IS who were followed up for two years, patients who received early statins within 28 days of their index stroke had a significantly lower risk of recurrent IS, mortality, ICH and other cardiovascular events compared to patients who did not. The present study adds to findings from previous studies, showing that the beneficial effects of early statins were still evident in relation to the elderly, across different ethnicities, and within the setting of varying statin dose intensity (low, moderate and high), statin lipophilicity (vs. hydrophilic), optimal LDL-C levels, LVO and embolic strokes, and different AF subtypes.

The observed benefits of early statin use in this cohort of AF patients with recent ischemic stroke can be attributed to a reduction in both cerebrovascular and cardiovascular events. Hence, statins may also play an important role in the secondary prevention of AF-related stroke.

In contrast to suggestions from prior studies, the present study found an association between early statin use and a reduction of recurrent stroke, with no increase in haemorrhagic stroke or ICH. It also confirms findings in previous studies that statins may reduce mortality and future cardiovascular events in patients with AF and recent stroke.

The pathophysiological basis for the beneficial effect of statins in IS remains unclear and a direct relationship between LDL-C and IS has not been firmly established. Nevertheless, clinical evidence for the preventive effects of statins in patients with a history of CHD, and those at risk of cardiovascular disease is clear^11,12,23–26^. The Stroke Prevention and Aggressive Reduction in Cholesterol Levels (SPARCL) trial established that high dose atorvastatin is effective for the prevention of recurrent IS in patients without CHD^13^. However, the results in the SPARCL study (and many other stroke secondary prevention trials^14–16^) cannot be generalised to all strokes (including AF-related stroke) because they specifically excluded patients who had AF or other cardioembolic sources of stroke. Nevertheless, numerous observational studies have shown that statins may reduce stroke recurrence, as well as improve survival and functional outcome in acute IS patients^27–29^. Although international practice guidelines follow the clinical evidence^17–20^, the position on use of post-stroke statins specifically in AF-related strokes remains unclear. Furthermore, patients were included in the SPARCL trial if they had suffered a stroke or TIA within the previous 1 to 6 months. Accordingly, the routine use of statins in the acute phase of stroke is not supported by strong evidence from any randomised clinical trials^30^.

Observational studies have shown that statin therapy is associated with reduced mortality and improved prognosis in IS patients with AF^31,32^ and patients with cardioembolic stroke^33,34^, and a lower risk of future cardiovascular events in cardioembolic^34^ and AF-related stroke patients^31,35^. However, there is limited evidence from those studies of any link between statin therapy and reduction of stroke recurrence. A significant association was not established despite a strong trend in favour of the group discharged on statins in one study involving patients with AF-related stroke^31^. More recently, a small prospective cohort study^36^ found that the use of statins may help to prevent stroke recurrence and improve functional outcomes in patients with cardioembolic stroke. In patients with embolic stroke of undetermined source, treatment with statins at discharge was associated with low rates of stroke recurrence, major adverse cardiovascular events and death^37^.

As well as a reduction of recurrent stroke risk, the present study suggests that statins may have a beneficial effect on reducing ICH following IS. This is at odds with the SPARCL trial where the reduction of recurrent stroke risk was counterbalanced by a small increase in the incidence of haemorrhagic stroke^13^. Such trend for increased haemorrhagic stroke has been found in other studies^38^ but not in the later Treat Stroke to Target clinical trial^39^. Although as a result of SPARCL, stroke practitioners are often reluctant to use statins following ICH, due to the uncertainty in the evidence, international guidelines have not imposed restrictions on the use of statins in such circumstances^19^. Indeed, the benefit of post-stroke statin therapy may outweigh its risks.

Given that clinical benefits of statins have been established in patients at high risk of cardiovascular disease and those with a history of heart disease, the influence of statins in the present study on reducing future stroke does not seem incongruous. As in the present study, patients with AF, as well as patients with a history of stroke, represent a population at high risk of atherosclerotic disease who often have concomitant comorbidities such as hypertension, CHD, heart failure or diabetes mellitus. A proportion of recurrent strokes in the present study may have been attributable to atherosclerotic causes. Our findings are aligned with the holistic or integrated care approach to AF management, including attention to comorbidities, and statins are clearly beneficial here^40,41^. This is important given the multimorbidity, frailty and inappropriate polypharmacy seen in patients with AF, with implications for treatments and outcomes^42–44^.

The mean age at index date of 73.7 years in our study was representative of that seen in the general population^45^. However, the incidence of stroke and post-stroke major adverse outcomes is high in the elderly^45^. Our sensitivity analyses in patients aged 75 years and over showed consistent results for all primary outcomes. In addition, LVO IS are more likely to be associated with AF^46^ and greater neurological deficits, and to contribute disproportionately to mortality and more severe functional outcomes after acute IS^47,48^. Consistent with the main analysis, our data suggest that statins may be associated with improved survival and cardiovascular outcomes even in patients with AF and recent LVO IS. Similar benefits were observed in patients with AF and recent embolic stroke.

When evaluating the effect of early statin use across different ethnic groups, we observed similar findings for the primary outcomes in the White ethnic group. In other ethnic groups studied (Black or African American, Hispanic, and Asian), we found an association between early statin use and reduced risk of recurrent IS and the composite outcome.

Stroke secondary prevention guidelines do not distinguish between AF subtype, and stroke risk is thought to be independent of AF subtype. Nevertheless, in IS patients, persistent AF is thought to be associated with a higher risk of recurrent IS^49,50^ and mortality^50^ than paroxysmal AF. Our data suggests that early statin use may have secondary prevention benefits in IS patients with either non-paroxysmal or paroxysmal AF, with beneficial effects on mortality in IS patients with paroxysmal AF.

Whether statins act in stroke only by lowering serum cholesterol or through effects beyond lipid lowering (so-called “pleiotropic effects”) remains a subject of debate. In the literature, the positive results in cardiovascular disease from clinical trials of proprotein convertase subtilisin/kexin type-9 (PCSK9) inhibitors and ezetimibe strengthen the argument that statins act through lipid-lowering^51,52^. PSCK9 inhibitors and ezetimibe are thought to act through lipid lowering, though the possible presence of other anti-thrombotic effects is emerging^53^.

However, as the trials were conducted on statin-treated patients, the position is not free from doubt, and elsewhere, anti-inflammatory interventions that do not act through lipid lowering, have been shown to reduce recurrent events compared with placebo^54^.

In the area of stroke, there is increasing evidence that statins act predominantly through lipid lowering^39,55–57^. The link between lipids and stroke is not as strong as the association between cholesterol levels and coronary artery disease, and appears to vary by stroke subtype^58–60^. The relationship seems strongest for atherosclerotic stroke subtypes, as supported by a recent meta-analysis of randomised clinical trials^59,61,62^. Nevertheless, the effect of statins in AF-related stroke remains unclear with most studies failing to demonstrate an association between dyslipidaemia and embolic stroke^60,63–65^.

In the present study, low, moderate and high intensity statins all reduced the risk of future stroke in the early statin group, which might suggest that the effects of statins in AF-related stroke are not only commensurate with the degree of lipid lowering. We also found that the risk of all primary outcomes were reduced in the early statin group even in patients with optimal levels of LDL-C less than 100 mg/dl. This might suggest that statins may have effects beyond lipid-lowering.

The pleiotropic effects of statins^66,67^ may extend to cardioprotective or neuroprotective effects that include changes in endothelial function, anti-inflammatory, anti-oxidant, anti-thrombotic activities (including reduced platelet activation), angiogenesis, and promotion of plaque stability^58,67^. Numerous experimental and animal models of stroke (including embolic stroke) support possible neuroprotective effects of statins^68–77^.

Although the finding of a lower 2-year risk of ICH in the early statin group in our study might be indirectly attributable to the non-lipid lowering properties of statins, a possible explanation could not be clearly derived in this study from the pleiotropic effects of statins.

Also, statins can be classified according to their synthesis (natural or synthetic), or their hydrophilicity. Lipophilic statins can more readily cross cell membranes and the blood brain barrier, and are thought to have greater pleiotropic effects (including neuroprotective effects on the central nervous system)^78^. In the present study, both lipophilic and hydrophilic statins reduced the risk of all primary outcomes in AF patients with recent IS. However, only lipophilic statins showed a significant reduction in the risk of ICH following stroke.

Nevertheless, as the 95%CI for the hydrophilic group likely exceeds 1 due to the smaller cohort and incidence of ICH, no strong conclusions can be drawn from this study regarding whether or not lipophilic statins may have stronger pleiotropic effects.

### Strengths and limitations

The main strengths of this study lie in the large size of the cohorts included in the analysis, and the use of propensity score matching to minimise the risk of bias from confounding.

Nevertheless, there were several limitations. Firstly, the retrospective nature of the study is more liable to error due to confounding and bias, and the observational nature cannot prove causality. Health care organization EMR data are subject to entry errors and data gaps, and some diagnoses may be underreported, while outcomes which occurred outside the studies network may have not been captured. The recording of ICD codes in EMR may vary by factors including age, comorbidities, severity of illness, length of in-hospital stay, and in-hospital mortality. Further residual confounding may include lifestyle factors such as alcohol consumption and physical activity, which were not available.

Use of a Cox model for analysis did not allow for adjustment for competing risks. This may have resulted in bias and overestimation of disease outcomes. Data were not available for cardiovascular and non-cardiovascular causes of mortality. This may have resulted in the overestimation of the absolute risk of all-cause mortality over time.

Assessments of disability following stroke using the modified Rankin Scale (mRS) or the National Institutes of Health Stroke Scale (NIHSS) scores were not available or insufficient. Accordingly, important information about patients’ stroke severity could not be determined from the available data, or used to support further analyses (including sensitivity analyses).

A degree of selection bias was likely present in the study because statins may not have been initiated in patients with severe neurological deficits and disabilities following stroke and those thought to have a poor prognosis. To minimise this, the study used propensity score matching and statistical adjustments to account for potential confounders.

The proportion of patients receiving early statins who later discontinued them, or the proportion of patients in the no-treatment group who later received statins is not known.

The study does not account for patients on combination therapy for dyslipidaemia, such as statin and ezetimibe. Data on underlying aetiologies of stroke were not available. Whether or not strokes were attributable to atherosclerosis, or embolic strokes resulted from emboli from the carotid arteries, aortic arch or the left ventricle, could not be determined. Finally, the effect of long term statin use was not investigated. An exploratory analysis excluding patients with history of cardiovascular disease was not conducted in the present study.

### Conclusions

Patients with AF and recent IS, who received statins within 28 days of their index stroke, had a lower 2-year risk of recurrent stroke, death, future cardiovascular outcomes and ICH, compared to those who did not.

## Data access and analysis

Dr Abdul-Rahim had full access to all of the data in the study and takes responsibility for the integrity of the data and the accuracy of the data analysis.

## Declaration of Interest

SEC,TB, JH, KHY, CTWT, AH and GI report no conflicts of interest.

KKL received grants from Croucher Foundation, Research Fund Secretariat of the Food and Health Bureau, Innovation and Technology Bureau, Research Grants Council, Amgen, Boehringer Ingelheim, Eisai and Pfizer; and consultation fees from Amgen, Boehringer Ingelheim, Daiichi Sankyo and Sanofi, all outside the submitted work.

GYHL reports Consultant and speaker for BMS/Pfizer, Boehringer Ingelheim, Daiichi-Sankyo, Anthos. No fees are received personally. GYHL is a National Institute for Health and Care Research Senior Investigator and co-principal investigator of the AFFIRMO project on multimorbidity in AF, which has received funding from the European Union’s Horizon 2020 research and innovation programme under grant agreement No 899871.

AHAR reports Member of the Editorial Board of Stroke and European Stroke Journal.

## Supplemental Material

Tables S1–S3

## Non-standard Abbreviations and Acronyms

AF: atrial fibrillation
CHD: coronary heart disease
EMR: electronic medical records
ESC: European Society of Cardiology
ESO: European Stroke Organisation
GDPR: General Data Protection Regulation
HCO: healthcare organisation
HIPAA: Health Insurance Portability and Accountability Act
ICD-10-CM: International Classification of Diseases, Tenth Revision, Clinical Modification
ICH: intracranial haemorrhage
LDL-C: low-density lipoprotein cholesterol
LVO: large vessel occlusion
MI: myocardial infarction
mRS: modified Rankin Scale
NIHSS: National Institutes of Health Stroke Scale
PCSK9: proprotein convertase subtilisin/kexin type-9
TIA: transient ischaemic attack

